# Background and Design of the Community Awareness Resources and Education (CARE) Project: Reducing Cervical Cancer in Appalachia

**DOI:** 10.1101/2020.04.01.20050211

**Authors:** Erinn M. Hade, Amy K. Ferketich, Amy M. Lehman, Electra D. Paskett, Mary Ellen Wewers, Mack Ruffin, Cathy Tatum, Stanley Lemeshow

## Abstract

**Objective:** The Community Awareness Resources and Education (CARE) project: Reducing Cervical Cancer in Appalachia is part of the NIH Centers for Population Health and Health Disparities initiative. It aims to increase screening rates for cervical cancer, to assist in tobacco cessation, and to determine the role of human papillomavirus (HPV) in contributing to the increased risk of cervical cancer.

**Methods:** Eligible subjects were recruited from 14 clinics in Appalachian Ohio. CARE’s cross sectional survey (CS) (571 participants) characterized social, behavioral, and environmental factors of Pap smear screening and smoking. The intervention projects tested programs to increase Pap smear screening (280 participants) or aid in smoking cessation (300 participants). A case control study (1360 participants) assessed social, behavioral, and biologic predictors of abnormal Pap tests.

**Results:** The CS participants tended to be younger, more educated, and were more likely to be employed than the general population of Appalachian women 18 and over. They were similar to the other samples in terms of race, marital status, income level and smoking status.

**Conclusions:** CARE will provide valuable information about the multi-level barriers to obtaining regular Pap smears and quitting smoking, as well as help to identify important biological and social risk factors related to abnormal Pap smears.

## Introduction

Cervical cancer is the second leading cause of cancer mortality among women worldwide (Boyle and Levin, 2008). The rates of cervical cancer incidence and mortality, however, have decreased by 75% since 1950 in the United States and other developed countries. The primary contributing factor for this decline is use of the Papanicolaou (Pap) smear screening test, which was introduced in the 1950s in the United States. Still, in 2008, an estimated 11,070 women in the U.S. were diagnosed with invasive cervical cancer and 3,870 women died from this cancer (American Cancer Society, 2008). Women in Appalachia have experienced a burden of excess cervical cancer incidence and mortality for many years. While HPV infection is often the ultimate cause of cervical cancer, four factors are believed to contribute to these elevated rates: 1) high prevalence of tobacco use (Prokopczyk et al., 1997, Simons et al., 1993, Simen-Kapeu et al., 2008); 2) low rates of Pap smear screening; 3) absence of proper and timely follow-up for abnormalities diagnosed from screening; and 4) survival differences due to lack of appropriate treatment (Schoenberg et al., 2005).

Links between cigarette smoking and cervical cancer were first observed in the late 1960s, with many subsequent investigations supporting this relationship (Daling et al., 1992, Giulano et al., 1997, Licciardone et al., 1989, Najuib et al., 1966, Prokopczyk et al., 1997, Winlelstein, 1990). Smoking prevalence in the Appalachian region of Ohio is estimated to be 28.2% among women, nearly 8% higher than the statewide estimate and 11% higher than the national estimate for women (American Cancer Society, 2007). Moreover, the rates among women of reproductive age are likely to be even higher (American Cancer Society, 2007). Past research suggests that cigarette smoking enhances immunosuppression, although the mechanisms behind this have not been fully determined (Castellsague and Munoz, 2003, Vineis et al., 2004, Giulano et al., 1997). Smoking may induce impaired antibody response in HPV16/18 infected young women, increasing risk for cervical cancer (Simen-Kapeu et al., 2008).

The Community Awareness Resources and Education (CARE) project is a P50 Center grant funded through the National Institutes of Health (NIH Centers for Population Health and Health Disparities initiative, RFA ES-02-009 (Paskett et al., 2008, Warnecke et al., 2008)). Consisting of both observational and intervention based projects, CARE uses a transdisciplinary approach to address the problem of high cervical cancer incidence and mortality rates in Ohio Appalachia (Holmes et al., 2008). The projects that are included in CARE are designed to gather multilevel information critical to improving the health of adult women in Appalachian Ohio, a region comprised of 29 counties in the state (Warnecke et al., 2008, Holmes et al., 2008, Paskett et al., 2008). The project aims are threefold: 1) increase screening rates for cervical cancer; 2) assist in tobacco cessation; and 3) determine the combined role of social/environmental, behavioral and HPV factors in contributing to the increased risk of cervical cancer.

To achieve these project aims, the following objectives are being examined: 1) identify social, environmental and behavioral barriers to obtaining regular Pap smears; 2) characterize social, behavioral and biological factors related to tobacco use; 3) evaluate a behavioral intervention to increase the rate of regular Pap smears; 4) test a tobacco cessation intervention; and 5) determine social/environmental, behavioral and biological factors that are associated with abnormal Pap test results.

The first phase of CARE, completed in June 2006, collected cross sectional (CS) in-person survey data on 571 women. Upon completion of the cross sectional interview, those among the 571 women who met the eligibility criteria were also invited to enroll in one of two intervention projects. The Pap Screening project (PS) tested an intervention to improve rates of Pap screening in this population. The Smoking Cessation project (SC) tested an intervention for helping women stop smoking. The third project, a case control study (CC), determined associations between social, behavioral and biological factors related to abnormal Pap test results. The purpose of this paper is to describe the design and background of the CARE study.

## Methods

Informed consent procedures and study protocols were reviewed and approved by the Institutional Review Boards of The Ohio State University, University of Michigan, and the Centers for Disease Control and Prevention.

## Study Population and Clinics

In order to represent women throughout the Ohio Appalachian region, counties were grouped into four regions. Each county was classified as urban or rural (using U.S. Census Bureau definitions) and within each region, two urban and two rural counties were selected and clinics were identified (see Figure 1). Counties in each region were randomly selected with probability proportional to the estimated average annual numbers of cervical cancer cases in each county from 1998-2000.

**Figure 1.**
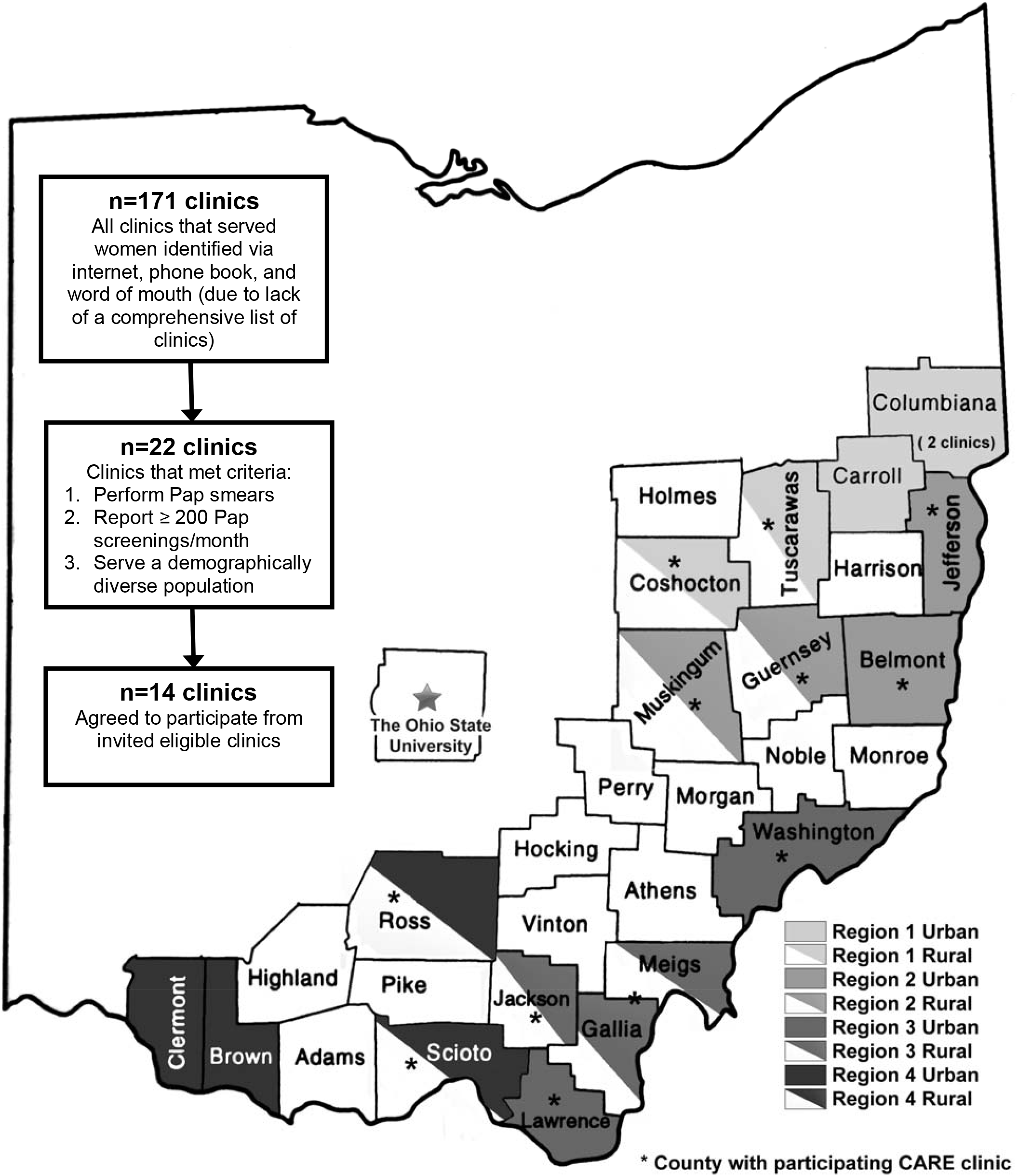
CARE County Selection, Appalachia Ohio 2005-2008.

## Cross Sectional Survey (CS)

The overall aim of the cross sectional survey was to characterize the social, behavioral and environmental factors related to Pap test screening within risk appropriate guidelines (Table 1) and tobacco use.

**Table 1.** Primary Outcomes/Risk Factors for the CARE Projects, Appalachia Ohio 2005-2008.

| **Project** | **Primary Outcomes** |
| --- | --- |
| **Cross Sectional Survey (CS)** | 1) Pap screening within risk appropriate guidelines (Cyrus-David et al., 2002, Hewitt et al., 2002):<br><u>*Risk Appropriate Guidelines Recommendations:*</u><br>*Annual screening for 'high risk' patients:*<br>• *Current smoker*<br>• *Had sexual intercourse at an early age*<br>• *Had a STD*<br>• *Have had a partner with a STD*<br>*Screening once every three years for 'low risk' patients:*<br>• *Patients that do not have any of the criteria for 'high' risk*<br>2) Current tobacco use |
| **Pap Screening Intervention (PS)** | 1) Whether or not a woman received a Pap smear screening between the date of randomization and the 12-month follow-up interview, as verified by medical record review |
| **Smoking Cessation Intervention (SC)** | 1) Cotinine-validated 7 day point prevalence abstinence at the 12 month follow up visit (Jarvis et al., 1987)<br>• Salivary cotinine levels $\leq 14\text{ng/mL}$ were considered to be non-smokers. |
| **Abnormal Pap Smear Case Control (CC)** | *Primary risk factors:*<br>1) Human papillomavirus (HPV) status<br>2) Presence of HPV oncogenic types<br>3) Serum HPV 16 antibody levels<br>4) First sexual intercourse before age 18<br>5) More than 4 sexual partners in lifetime<br>6) Current smoker<br>7) Pap test screening (in last year, last 3 years, or ever). |

### Screening and Recruitment

Women were randomly sampled from listings of all eligible women (eligibility criteria listed in Table 2) from each of the 14 study clinics (Figure 1). Sampling of potential participants occurred on a monthly basis and sample lists were updated every six to eight months in order to capture newly eligible patients. Following the initial eligibility screening of sampled medical records, patients were sent a letter inviting them to participate. Women who did not return postcards accepting or declining participation were contacted by phone. At least 10 attempts were made to contact potential participants by phone to evaluate eligibility. Those women who were eligible and agreed to participate were contacted by a study interviewer, who then obtained written consent and conducted the CS interview.

**Table 2.**
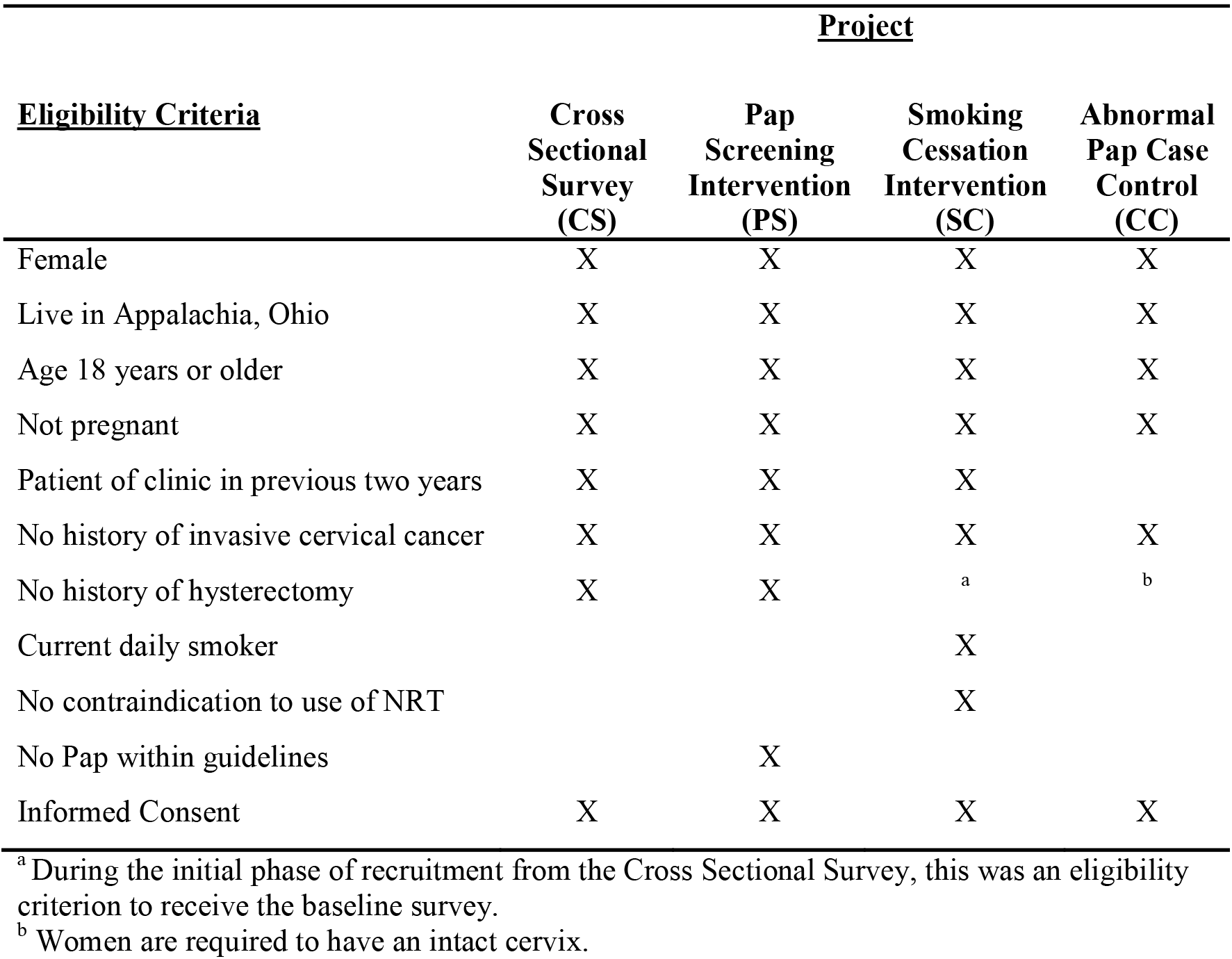
CARE eligibility criteria by project, Appalachia Ohio 2005-2008.

### Timeline, Measures, and Outcomes

Recruitment began in March of 2005 and was completed in June 2006 for the CS study, with 571 participants. Interview measures are outlined in Table 3; the main outcomes included Pap screening within risk appropriate guidelines and current tobacco use (Table 2) (Cyrus-David et al., 2002, Hewitt et al., 2002). In-person CS interviews were administered using a Computer Assisted Personal Interview (CAPI) system, which included an audio portion to collect sensitive data.

**Table 3.**
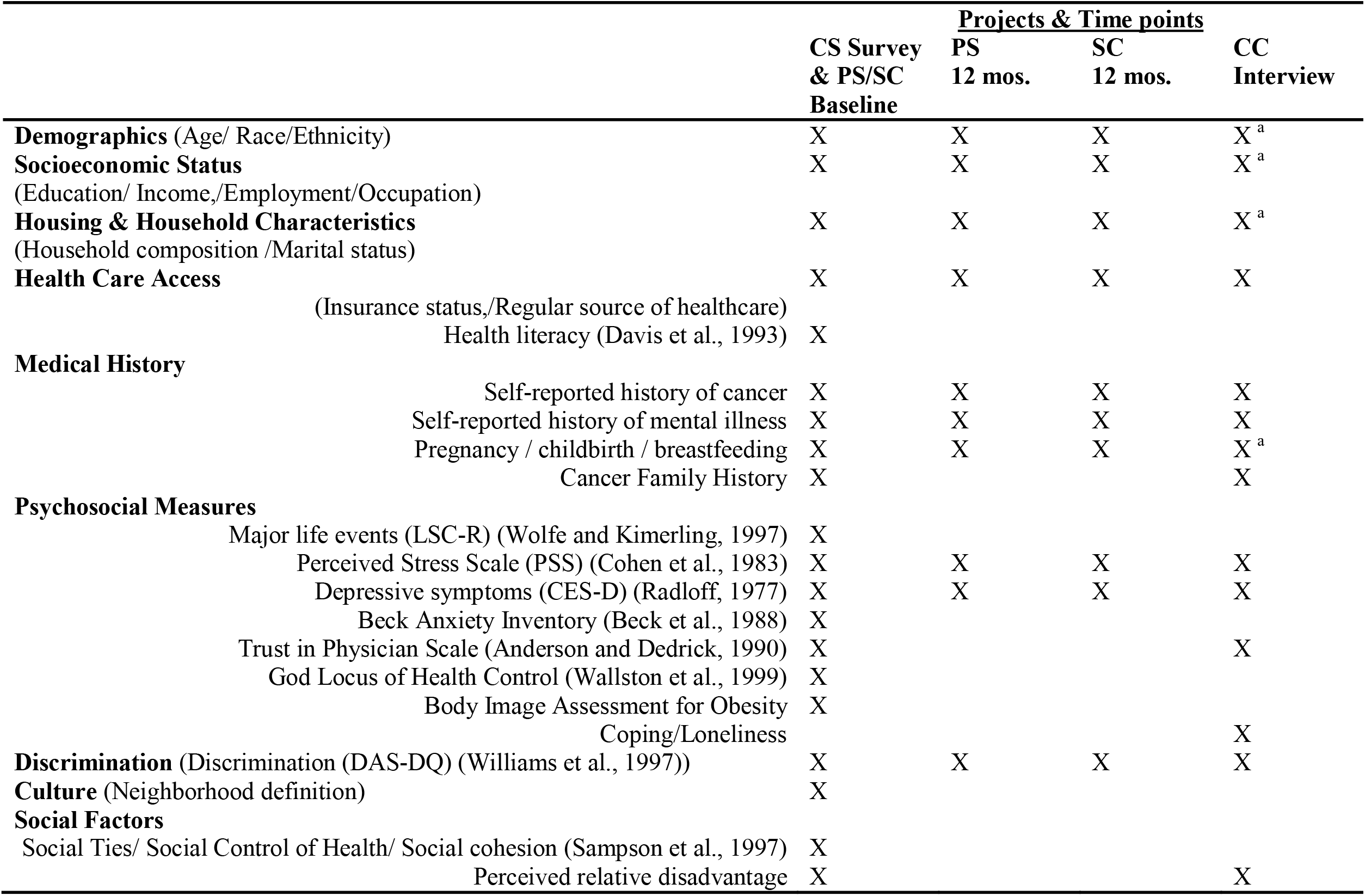
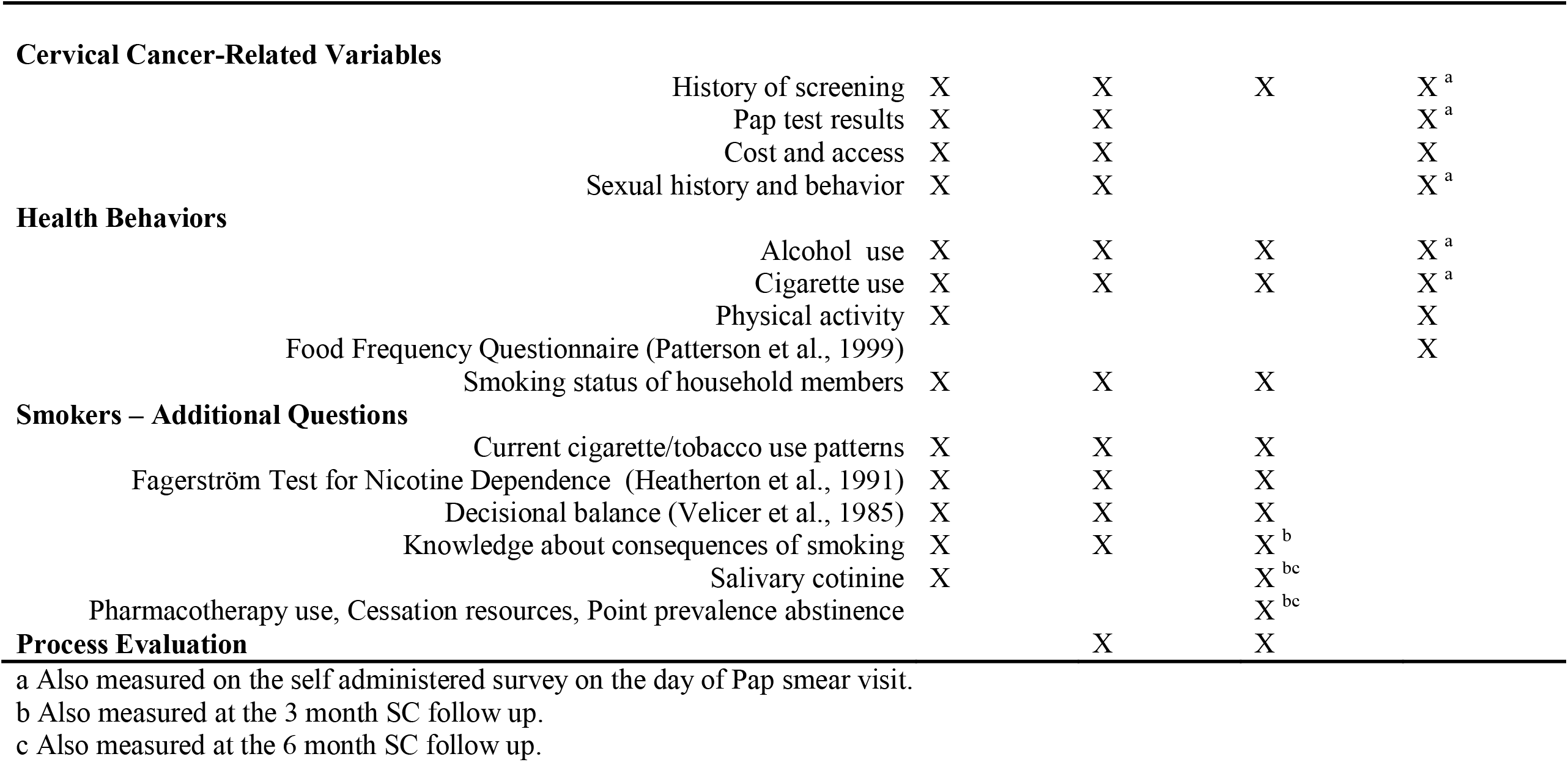
CARE measures by data collection period, Appalachia Ohio 2005-2008.

|  | CS Survey<br>& PS/SC<br>Baseline | Projects & Time points |  |  |
| --- | --- | --- | --- | --- |
|  |  | PS<br>12 mos. | SC<br>12 mos. | CC<br>Interview |
| **Demographics** (Age/ Race/Ethnicity) | X | X | X | X<sup>a</sup> |
| **Socioeconomic Status**<br>(Education/ Income./Employment/Occupation) | X | X | X | X<sup>a</sup> |
| **Housing & Household Characteristics**<br>(Household composition /Marital status) | X | X | X | X<sup>a</sup> |
| **Health Care Access**<br>(Insurance status./Regular source of healthcare) | X | X | X | X |
| Health literacy (Davis et al., 1993) | X |  |  |  |
| **Medical History** |  |  |  |  |
| Self-reported history of cancer | X | X | X | X |
| Self-reported history of mental illness | X | X | X | X |
| Pregnancy / childbirth / breastfeeding | X | X | X | X<sup>a</sup> |
| Cancer Family History | X |  |  | X |
| **Psychosocial Measures** |  |  |  |  |
| Major life events (LSC-R) (Wolfe and Kimerling, 1997) | X |  |  |  |
| Perceived Stress Scale (PSS) (Cohen et al., 1983) | X | X | X | X |
| Depressive symptoms (CES-D) (Radloff, 1977) | X | X | X | X |
| Beck Anxiety Inventory (Beck et al., 1988) | X |  |  |  |
| Trust in Physician Scale (Anderson and Dedrick, 1990) | X |  |  | X |
| God Locus of Health Control (Wallston et al., 1999) | X |  |  |  |
| Body Image Assessment for Obesity | X |  |  |  |
| Coping/Loneliness |  |  |  | X |
| **Discrimination** (Discrimination (DAS-DQ) (Williams et al., 1997)) | X | X | X | X |
| **Culture** (Neighborhood definition) | X |  |  |  |
| **Social Factors** |  |  |  |  |
| Social Ties/ Social Control of Health/ Social cohesion (Sampson et al., 1997) | X |  |  |  |
| Perceived relative disadvantage | X |  |  | X |
| **Cervical Cancer-Related Variables** |  |  |  |  |
| History of screening | X | X | X | X<sup>a</sup> |
| Pap test results | X | X |  | X<sup>a</sup> |
| Cost and access | X | X |  | X |
| Sexual history and behavior | X | X |  | X<sup>a</sup> |
| **Health Behaviors** |  |  |  |  |
| Alcohol use | X | X | X | X<sup>a</sup> |
| Cigarette use | X | X | X | X<sup>a</sup> |
| Physical activity | X |  |  | X |
| Food Frequency Questionnaire (Patterson et al., 1999) |  |  |  | X |
| Smoking status of household members | X | X | X |  |
| **Smokers – Additional Questions** |  |  |  |  |
| Current cigarette/tobacco use patterns | X | X | X |  |
| Fagerström Test for Nicotine Dependence (Heatherton et al., 1991) | X | X | X |  |
| Decisional balance (Velicer et al., 1985) | X | X | X |  |
| Knowledge about consequences of smoking | X | X | X<sup>b</sup> |  |
| Salivary cotinine | X |  | X<sup>bc</sup> |  |
| Pharmacotherapy use, Cessation resources, Point prevalence abstinence |  |  | X<sup>bc</sup> |  |
| **Process Evaluation** |  | X | X |  |

a Also measured on the self administered survey on the day of Pap smear visit.
b Also measured at the 3 month SC follow up.
c Also measured at the 6 month SC follow up.

### Sample Size

The sample size for the CS was based on the number of interviews needed to estimate the prevalence of Pap screening (within guidelines) and current smoking. With a margin of error set to 5%, assuming that simple random sampling would be performed with a population size equal to 15,000, we estimated that at least 415 women were needed to estimate risk appropriate Pap screening and current smoking prevalence with the given precision.

### Participant Flow

Figure 2 details participant recruitment into the CS survey. Potential participants were sent a letter that described the study and specified they would be called by study staff. Of those sampled and initially reviewed for eligibility, 1754 subjects were contacted, resulting in 1038 potentially eligible subjects. Subsequently, 237 of these subjects were deemed ineligible or could not be contacted, leaving 801 subjects who were eligible and able to be contacted. The participation rate in the CARE CS survey was 71%. As shown in Table 4, the response rate was 28% (The American Association for Public Opinion Research 2008).

**Table 4.** CARE Cross Sectional Survey Patient Recruitment, Appalachia Ohio 2005-2006.

|  |  |
| --- | --- |
| **A. Subjects interviewed:** | **571** |
| **B. Subjects who were sampled and known to be eligible but refused or unable to participate:** | **424** |
| Eligible subjects contacted, refused before the interview | 86 |
| Eligible subjects contacted, refused at the interview | 144 |
| Eligible subjects unable to be contacted after agreeing to the study | 194 |
| **C. Subjects who were sampled with eligibility status unknown:** | **1728** |
| Subjects who could not be contacted after initial chart review | 1051 |
| Subjects who refused after being contacted | 579 |
| Subjects who mailed in a refusal | 98 |
| **D. Estimated number of eligible subjects among those sampled without known eligibility status (approximately 60% of C):** | **1030** |
| **E. Calculated total number of eligible subjects (A + B + D):** | **2025** |
| **F. Estimated Response Rate (A/E):** | **28%** |
| **G. Subjects who were eligible and agreed to be contacted (from Figure 2):** | **801** |
| **H. Estimated Participation Rate (A/G):** | **71%** |

**Figure 2.**
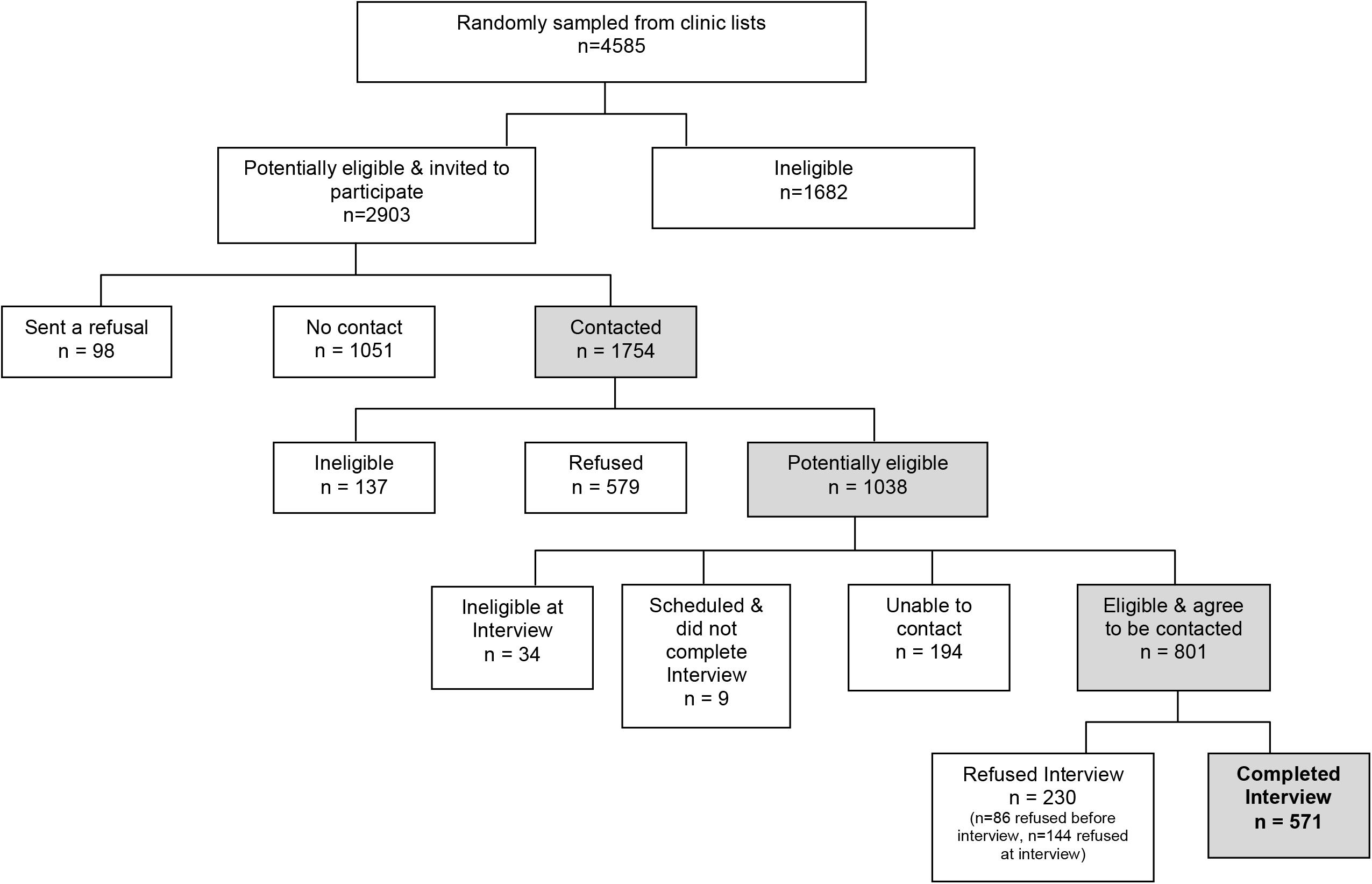
CARE Cross Sectional Survey Patient Flow, Appalachia Ohio 2005-2006.

**Figure 3.**
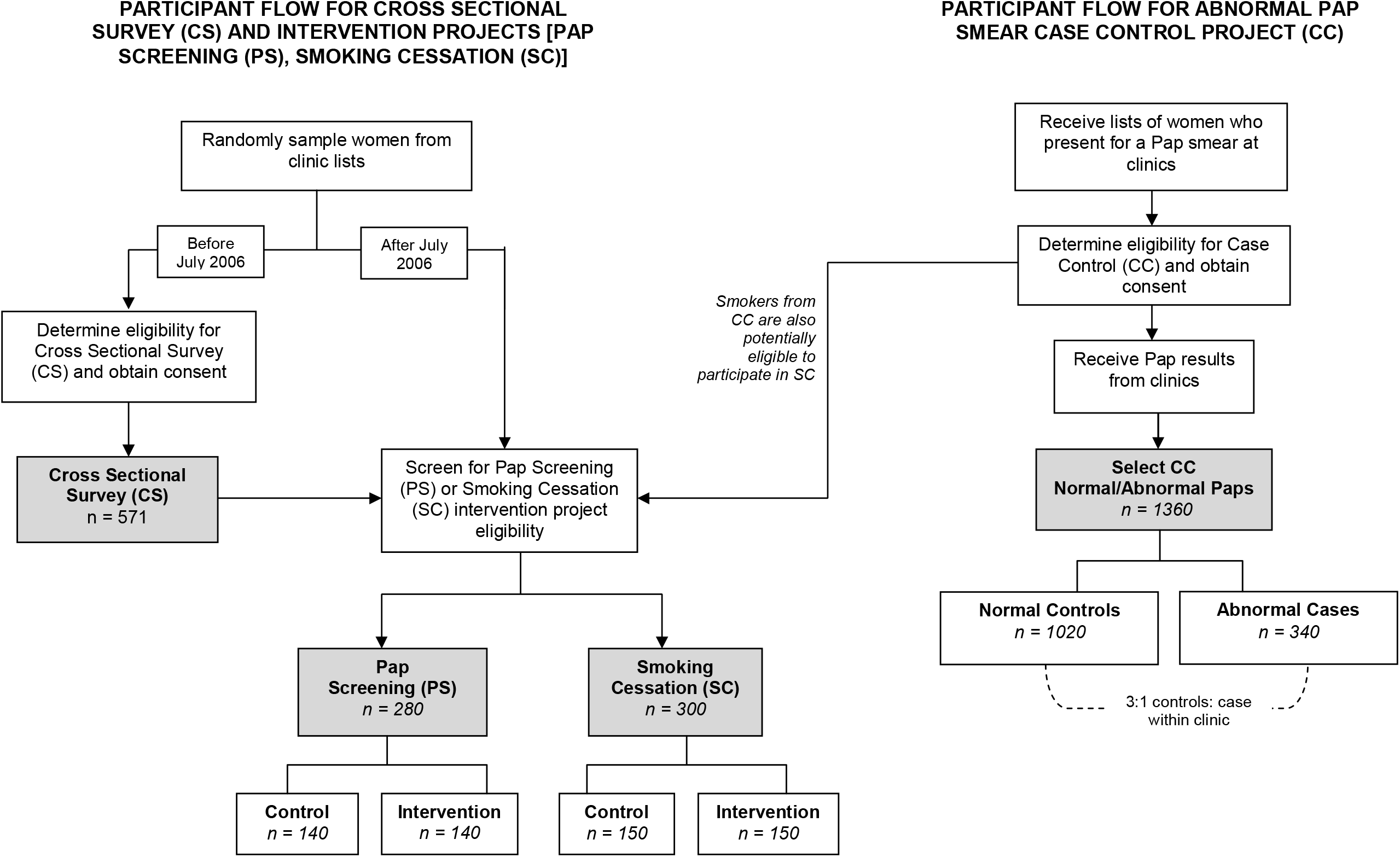
CARE Cross Sectional Survey Participants and Target Sample Sizes for the Intervention and CC Projects, Appalachia Ohio, 2005-2008.

### Intervention Studies

The Pap screening (PS) and smoking cessation (SC) intervention studies used similar methods; thus they are presented together below.

#### Recruitment and Screening

Eligible participants (criteria listed in Table 2) were initially recruited from the 571 individuals who completed the cross sectional survey; following the completion of the CS in July 2006, participants were identified as described above, contacted by letter and then screened via the phone for eligibility (Figure 3). In addition to the screening process, smokers who completed participation in the case control (CC) project were also recruited into the SC project. All eligible women who were consented received a baseline interview prior to randomization.

#### Interventions

Both interventions included two arms: a usual-care control group that received information by mail and an intervention group that received visits by a lay health advisor (LHA) (Table 5). The LHAs were women from the same community as the participants who were trained to educate women on the importance of having a routine Pap test or quitting smoking.

**Table 5.** Description of Treatment Arms for the CARE PS and SC Interventions, Appalachia Ohio 2005-2008.

| **Intervention Project** | **Control** | **Intervention** |
| --- | --- | --- |
| **Pap Screening Intervention (PS)** | 1) Send letter from woman's physician encouraging her to have a Pap test along with NCI brochure about the importance of Pap tests | 1) Initial visit from LHA<br>2) *1 month after initial visit*: Telephone call from LHA<br>3) *2 months*: Postcard<br>4) *3 months*: Postcard<br>5) *5 months*: Telephone call from LHA<br>6) *6 months*: Postcard<br>7) *7 months*: Postcard<br>8) *10 months after initial visit*: Second visit from LHA |
| **Smoking Cessation Intervention (SC)** | 1) Send letter from physician at woman's medical clinic encouraging smoking cessation (Fiore et al., 2000) along with self-help informational sheet designed by the U.S. Public Health Service entitled *You Can Quit Smoking Consumer Guide* (Fiore et al., 2000) | 1) *Week 1*: Initial visit from LHA<br>2) *Week 2*: Visit 2, set quit date<br>3) *Week 3*: Visit 3, receive NRT patches<br>4) *Week 4*: Visit 4<br>5) *Week 6*: Visit 5<br>6) *Week 8*: Visit 6<br>7) *Week 10*: Visit 7 |

Participants enrolled in the intervention arm of PS received two visits from the LHA. The LHAs visited women either in their home or at a site in the community. During the visit, the LHA conveyed messages designed to increase knowledge about cervical cancer, Pap test screening, and the importance of follow-up after an abnormal test. The LHA also assessed each woman’s personal risk of getting cervical cancer in her lifetime and provided individualized counseling for reported barriers to having a Pap test. The LHA maintained contact with the participant following the initial visit with telephone calls and postcards (Table 5), culminating in a second visit approximately 10 months after the first. At the final visit, the LHA provided additional barriers counseling, if necessary, and encouraged the woman to continue to be proactive about her healthcare. Women in the control group received a letter from their physician after the baseline interview that encouraged them to have a Pap test, along with an NCI brochure about the importance of Pap tests.

Women in the SC project received seven visits by the LHA over a 10-week period (Table 5). Participants set a quit date during visit two and received nicotine replacement therapy (NRT) patches at visit three. In visits four through seven the LHAs assessed slips and NRT side effects, promoted cessation to women who had not quit, congratulated women who quit and encouraged continued cessation, delivered additional NRT patches, and collected any unused NRT patches. At each visit, the LHA collected participant data including current medications and medication changes. All symptoms associated with NRT were reported to a nurse who case managed patients through the LHAs. Women in the control group received a letter from the physician at their medical clinic that encouraged smoking cessation and a self-help informational sheet designed by the U.S. Public Health Service entitled *You Can Quit Smoking Consumer Guide* (Fiore et al., 2000).

#### Timeline, Measures and Outcomes

Recruitment into the intervention projects continued through January 2008. The primary outcome for the PS project was whether or not a woman received a Pap smear screening between the date of randomization and the 12-month follow-up interview, as verified by medical record review. In addition, self-reported Pap smear information was collected.

For the SC project, follow-up interviews were conducted at 3, 6, and 12 months following randomization. The primary outcome was cotinine-validated 7 day point prevalence abstinence at the 12 month follow up visit (Jarvis et al., 1987). Additional outcomes included quit rates at 3 and 6 months, prolonged abstinence and the number of serious quit attempts. The baseline interview collected information on a variety of topics, similar to that in the CS survey.

Follow-up of participants in the intervention projects continued through the spring of 2009. Data verification and processing is ongoing as of the summer of 2009.

## Abnormal Pap Smear Case Control Project (CC)

Although risk factors have been identified for cervical cancer, with HPV infection of primary importance, it is unclear which of these risk factors contributes to disparity in cervical cancer incidence and mortality seen in Appalachia. The CC component of CARE seeks to determine social, behavioral and biological risk factors in this Appalachian Ohio population and to determine which combination of these factors contribute to developing cervical cancer. Moreover, the CC project will provide some of the first estimates of HPV infection, oncogenic HPV types and variants to HPV in Appalachian women.

### Screening and Recruitment

Women who met eligibility criteria (Table 2) were invited to participate in the CC project during a scheduled Pap smear at one of the study clinics. Study nurses recruited women who had scheduled a Pap smear at one of the CARE participating clinics. At that time potential participants signed a written informed consent, completed a short self-administered questionnaire, and provided blood and saliva samples (Figure 3). An additional smear was taken and a sample for HPV typing was obtained by the physician.

Cases were defined to be women with an abnormal Pap smear according to the 2001 Bethesda System for Reporting Pap Smear Results, including abnormalities from atypical squamous cells of uncertain significance (ASCUS) to cervical cancer(Solomon et al., 2002). For each case, three controls were randomly sampled from the same clinic as the case among recruited patients who had normal Pap results. All controls were selected within a three month window around the time the case received her Pap smear.

### Timeline, Measures, and Outcomes

Items obtained from the self-administered survey and the telephone CC interview are listed in Table 3. The self-administered survey contained questions regarding demographics, Pap smear test history, reproductive health and sexual activity history, as well as alcohol and tobacco use. Prior to June 2008 a telephone interview was collected for cases and controls after the cytology review was complete. Thereafter, the interview was given at the time of recruitment. HPV was determined using the commercially manufactured PCR-based Roche AMPLICOR (Stevens et al., 2007, Schalasta et al., 2007, Monsonegoa et al., 2005). Serum antibody levels were determined using a HPV multiplexed competitive Luminex immunoassay (Opalka et al., 2003, Dias et al., 2005). Recruitment of participants ended in December 2008 and sample processing and cleaning are ongoing as of the summer of 2009.

### Sample size

Sample size estimates were calculated for seven primary risk factors (Table 1) assuming a two-sided chi-square test, type I error rate of 0.05 and 80% power. The largest required sample size of 1044 (261 cases and 783 control subjects) was calculated for comparing the proportion of HPV positive subjects between cases (0.55) and controls (0.45). We estimated that sampling approximately 340 cases and 1020 controls would provide adequate power given some attrition/missing survey data and inadequate samples for processing.

## Results

### Baseline Characteristics in Cross Sectional Survey

Demographics of the CS participants are presented in Table 6. Most participants were younger than 65, were white, had at least a high school diploma or GED, and had health insurance. As expected, just over 27% of our sample reported being current smokers.

**Table 6.** CARE Cross Sectional Survey participant characteristics versus those of Appalachian women from other data sources, Appalachia Ohio 2005-2006 [*% (95% CI)*]

| *Characteristic* | *CARE CS<sup>a</sup>* | *BRFSS<sup>b</sup>* | *ATS<sup>c</sup> (n=575)* | *OFHS<sup>d</sup>* | *Census<sup>e</sup>* |
| --- | --- | --- | --- | --- | --- |
| % under age 65 | 95.3 | 77.7 (72.3-83.2) | 79.9 (75.3-84.5) | 78.4 (76.8-80.1) | 84.2 |
| % with ≥ HS diploma (among women ≥ 25 yrs) | 92.5 | 86.7 (82.1-91.3) | 89.3 (85.9-92.7) | 85.9 (84.5-87.3) | 78.5 |
| % White | 94.7 | 92.3 (88.6-96.1) | 94.5 (91.5-97.4) | 96.3 (95.6-97.0) | 97.0<sup>f</sup> |
| % Married | 61.1 | 67.4 (60.9-73.8) | 64.0 (57.9-70.1) | 58.5 (56.6-60.4) | 58.0 |
| % with PIR < 1 | 25.6 | NA | NA | 23.1 (21.4-24.8) | 13.6<sup>g</sup> |
| % Currently employed | 68.7 | 46.3 (39.2-53.4) | 46.6 (40.2-53.0) | 46.4 (44.5-48.4) | 49.5 |
| % With income < \$25,000 | 42.9 | 39.9 (32.6-47.2) | 38.8 (31.8-45.7) | NA | NA |
| % With no health insurance | 15.2 | 20.3 (14.0-26.6) | 17.2 (11.8-22.5) | 14.2 (12.8-15.5) | NA |
| % Current smokers | 27.4 | 27.7 (21.0-34.4) | 28.2 (22.4-33.9) | 28.2 (26.4-29.9) | NA |

<sup>a</sup>CARE Cross Sectional survey data collected in 2005 and 2006
<sup>b</sup>BRFSS data collected in 2005 (Appalachian stratum only in estimate)
<sup>c</sup>Ohio Adult Tobacco Survey (ATS) data collected in 2005 and 2006 (Appalachian stratum only in estimate)
<sup>d</sup>OFHS data collected in 2004 (Appalachian counties only in estimate and only women who have been to provider in past 2 years)
<sup>e</sup>Census 2000 data collected (Appalachian counties only in estimate)
<sup>f</sup>Census 2000 estimate also includes men and is calculated among individuals who reported one race
<sup>g</sup>Census 2000 estimate also includes men (no separate data for women)
NA = Not Available

In order to gauge how comparable our CS participants were to the population of adult women in Appalachian Ohio, we compared demographic characteristics to a number of data sources that describe this population (Table 6). Data sources included the Behavioral Risk Factor Surveillance System 2005 (BRFSS) (Centers for Disease Control and Prevention (CDC), 2005), the Ohio Adult Tobacco Survey 2005-2006 (ATS) (Renaud et al., 2006), the Ohio Family Health Survey 2004 (OFHS) (State of Ohio Department of Job and Family Services, 2005) and the 2000 Census (U.S. Census Bureau, 2001). In comparison to other samples, the CS participants tended to be younger, more educated, and were more likely to be employed than the general population of women 18 and over in Appalachian Ohio. However, the CS participants were similar to the other samples in terms of race, marital status, income level and smoking status.

## Discussion

Previous research in Pap test screening and tobacco cessation in communities with health disparities has been reported, however none has focused on women in Appalachian Ohio. Similar intervention strategies involving LHA’s tailored to community specific barriers have been reported for mammography screening in underserved populations from North Carolina (Paskett et al., 2006, Paskett et al., 2004, Paskett et al., 1999). However these studies differed in their overall design; focusing on different screening programs and including fewer clinics. CARE has involved 14 clinics from throughout the region, aiming to provide a broader picture of intervention effects across the Appalachian region of Ohio.

Several tobacco cessation intervention programs have been implemented in rural Ohio (Wewers et al., 2002, Wewers et al., 2006). These differed from the current study in several ways: they did not focus on female smokers; they implemented county wide intervention or control protocols; and they recruited participants from clinics rather than sampling potential participants. The smoking cessation intervention of CARE includes a local LHA and management of the tobacco treatment by a nurse. Comparison of recruitment and retention as well as treatment effects between the studies will contribute to our understanding of successful tobacco intervention programs.

Given that our recruitment strategy was clinic based, it was not surprising that there were demographic differences between the CS sample and that of the general population of Appalachian women. For the population of women who visited clinics, we could approximate their demographic characteristics from the OFHS (women who indicated that they have visited a doctor in the past two years). While the OFHS had limitations associated with a phone survey, our sample was still younger, more educated and more likely to have insurance than those women in OFHS who said that they had seen a physician recently. Further description of the CS participants can be found in forthcoming articles on determinants of Pap smear testing within guidelines and on smoking status (Paskett et al., Accepted, Wewers et al., Under review).

As has been a common challenge of survey and community based research, recruitment of participants into each of the studies, observational and intervention alike, has been an ongoing challenge in CARE. Difficulties in recruitment have stemmed more from non-contact of many potentially eligible participants and less from refusal to participate. Curtin, Presser and Singer report a sharp decline in response rates from 1996 to 2003 in a study spanning from 1979 through 2003 (Curtin et al., 2005). In the past twenty years, non-contact rates have grown to reach similar levels as refusal rates, and the authors speculated that this is consistent with the increased use of caller id (Curtin et al., 2005). This argument is consistent with the experience of CARE investigators.

## Conclusion

CARE is unique in the CPHHD initiative in that it is the only Center that focuses on a rural population. The observational and intervention projects that comprise CARE will provide valuable multilevel information about the environmental and behavioral barriers to obtaining regular Pap smears, as well as help to identify important biological (HPV) and social risk factors related to the underlying mechanisms of abnormal Pap smears. Moreover, CARE will provide important information about social and behavioral characteristics of tobacco use among women in Appalachian Ohio and the effectiveness of lay-health interventions for smoking cessation and cervical cancer screening will be tested in this particular population.

## Data Availability

Data can be made available upon request to the study PIs.

## Acknowledgements

This study was supported by National Institutes of Health Grant P50CA105632 and the Behavioral Measurement Shared Resource at The Ohio State University Comprehensive Cancer Center, Grant P30CA016058 from the National Cancer Institute.

